# Postprandial glycaemic response in different ethnic groups in East London and its association with vitamin D status: study protocol for an acute randomised crossover trial

**DOI:** 10.1101/2024.11.28.24318133

**Authors:** Honglin Dong, Christian Reynolds, AFM Saiful Islam, Swrajit Sarkar, Sophie Turner

## Abstract

In the UK, black African-Caribbeans (ACs) and South Asians (SAs) have 3–6 times greater risks of developing diabetes than white Caucasians do. East London is among the areas with the highest prevalence of type 2 diabetes and the highest proportion of minority groups. This ethnic health inequality is ascribed to socioeconomic standing, dietary habits, culture, and attitudes, while biological diversity has rarely been investigated. The evidence shows that the postprandial glucose peak values in SAs are 2–3 times greater than those in white Caucasians after the same carbohydrate loads; however, the mechanism is poorly understood. In the UK, 50% of SAs and 33% of ACs have vitamin D (vitD) deficiency, whereas 18% of white Caucasians have vitamin D deficiency. There is evidence that vitD status is inversely associated with insulin resistance in healthy adults and diabetic patients and that vitD supplementation may help improve glycaemic control and insulin resistance in type 2 diabetes patients. However, little evidence is available on minority groups or East London. This study will investigate the postprandial glycaemic response (PGR) in three ethnic groups (white Caucasians, SAs and ACs) in East London and link PGR to plasma 25(OH)D (an indicator of vitD status). Ninety-six healthy adults (n=32 per group) will be recruited. Two test drinks will be provided to the participants (300 ml of glucose drink containing 75 g glucose, and 300 ml of pure orange juice) on different occasions. PGR is monitored before and after drinking every 30 min for up to 2 hours via finger prick. A fasting blood sample obtained via phlebotomy will be used for 25(OH)D and relevant tests. A knowledge/perception questionnaire about vitD and a 4-day food diary (analysing vitD dietary intake) will also be collected. The findings of the study will be shared with participants, published in journals, disseminated via social media, and used to inform a randomized controlled trial of the effects of vitD supplementation on PGR in minority groups.

The study complies with the Helsinki Declaration II and was approved by the Senate Research Ethics Committee at City, University of London (ETH2223-2000). The study findings will be published in open access peer-reviewed journals and disseminated at national and international conferences. ClinicalTrials.gov Identifier: NCT06241976

## Introduction

Background Health patterns differ significantly between ethnic minority groups and the white population. In the UK, the risk of developing diabetes is 3-6 times greater in South Asians (SAs) and up to three times greater in black African-Caribbeans (ACs) than in white Caucasians, and people in these groups develop this condition at a younger age^(1)^. East London is among the areas with the highest proportion of minority groups^(2)^ and the highest prevalence of type 2 diabetes mellitus (T2DM)^(3)^. Although multiple factors, including socioeconomic standing, diet, culture and attitudes, language barriers, genetics and lifestyles, have been identified^(4)^, research into biological diversity is scarce. Recent research revealed that the postprandial glucose peak in SAs is two- to three-fold greater than that in white Caucasians after identical carbohydrate loads are reached^(5)^. Although obesity is believed to account for 80–85% of the risk of developing T2DM due to obesity causing insulin resistance^(6)^ and some minority groups, e.g., black people, have a higher prevalence of overweight and obesity than white British people do (73.6% vs. 63.3%)^(7)^, other biological mechanisms, including vitamin D (vitD) deficiency, are poorly understood.

VitD deficiency in minorities in the UK is well known and is described as an unrecognised epidemic^(8)^. In the UK, 50% of SAs and 33% of black ACs demonstrate vitD deficiency, whereas 17.5% of white Caucasians do^(9)^, which is primarily due to more subcutaneous pigmentation that absorbs ultraviolet B from sunlight and reduces vitD production in the skin and at high latitudes in the UK^(10)^. This situation is worse in East London. In Tower Hamlets, a borough of East London, 47% of black and 42% of Asians have vitD deficiency, whereas 17% of the white population has such deficiency^(11)^. An inverse association of serum 25(OH)D levels with insulin resistance was observed in healthy adults^(12)^ and diabetic patients^(13)^. Recent evidence has shown that vitD supplementation may help improve glycaemic control and insulin resistance in T2DM patients^(14)^. However, little evidence is available for minority groups or residents in East London, indicating that AC and SA communities are underrepresented in the evidence base concerning diabetes and vitamin D. VitD plays important roles in calcium metabolism and is involved in the modulation of cell growth, neuromuscular and immune function, and the reduction of inflammation due to its receptors being expressed ubiquitously in nearly all human cells, including pancreatic β-cells^(15)^. Animal studies have shown that vitD treatment improves insulin production and sensitivity^(16)^, and increased insulin secretion may be caused by increased intracellular calcium(17). Moreover, 1,25(OH)2D (the active form of vitD) may modulate β-cell growth and differentiation^(15)^. The secondary high parathyroid hormone (PTH) concentration^(18)^ and increased inflammatory markers^(19)^ associated with vitD deficiency may also cause glucose intolerance. VitD may have an indirect effect on glycaemic control via obesity. Our research (accepted for publication, attached) and many others^(20)^ revealed a significant inverse association between body mass index (BMI) and serum 25(OH)D, which is thought to involve a complex of mutual influences because vitD receptors are expressed on adipose cells and regulate their functions^(21)^, indicating that vitD deficiency might be one of the causes of obesity, thus indirectly leading to an increased risk of T2DM.

The postprandial glycaemic response (PGR) has implications for T2DM development^(22)^. The oral glucose tolerance test (OGTT) is widely used to assess insulin sensitivity and pancreatic β-cell function and to assess an individual’s metabolic capacity to handle carbohydrate-containing foods^(23)^. However, a recently published study indicated that within-subject variations in the PGR pattern may exist between OGTT and food intake, suggesting the necessity of combining OGTT and a meal/drink tolerance test for individualized glycaemic management^(24)^. The awareness of vitD and its impact on health is poor in the UK. Although the COVID-19 pandemic has attracted the attention of the public on vitD and, a recent UK survey^(25)^ revealed that 49% of adults are unaware of the UK government’s guidelines for vitD. There is no such survey available on minority groups or residents in East London. We are also interested in dietary vitD intake between different ethical groups in East London, which will partly explain the vitD status of the target population. There is an urgent call for research on minority populations to address health inequality^(26)^. This proposal is an attempt to respond to the above call with a focus on minority communities in East London.

The aims of this study were to investigate the differences in PGR to OGTT and OJ consumption among white Caucasian, SA and AC adults; to investigate the associations of the plasma 25(OH)D concentration with PGR to OGTT and OJ consumption in white Caucasian, SA and AC adults; and to assess the knowledge and perception of vitD and dietary vitD intake in white Caucasian, SA and AC adults.

## Method

This is an acute randomised, repeated measures crossover trial. Figure 1 shows the study flow chart. The study was approved by the Senate Research Ethics Committee at City, University of London (**ETH2223-2000**). The recruitment period of the study is between 1^st^ November 2023 and 31^st^ December 2024. All participants gave written consent before taking part in the study.

**Figure 1.**
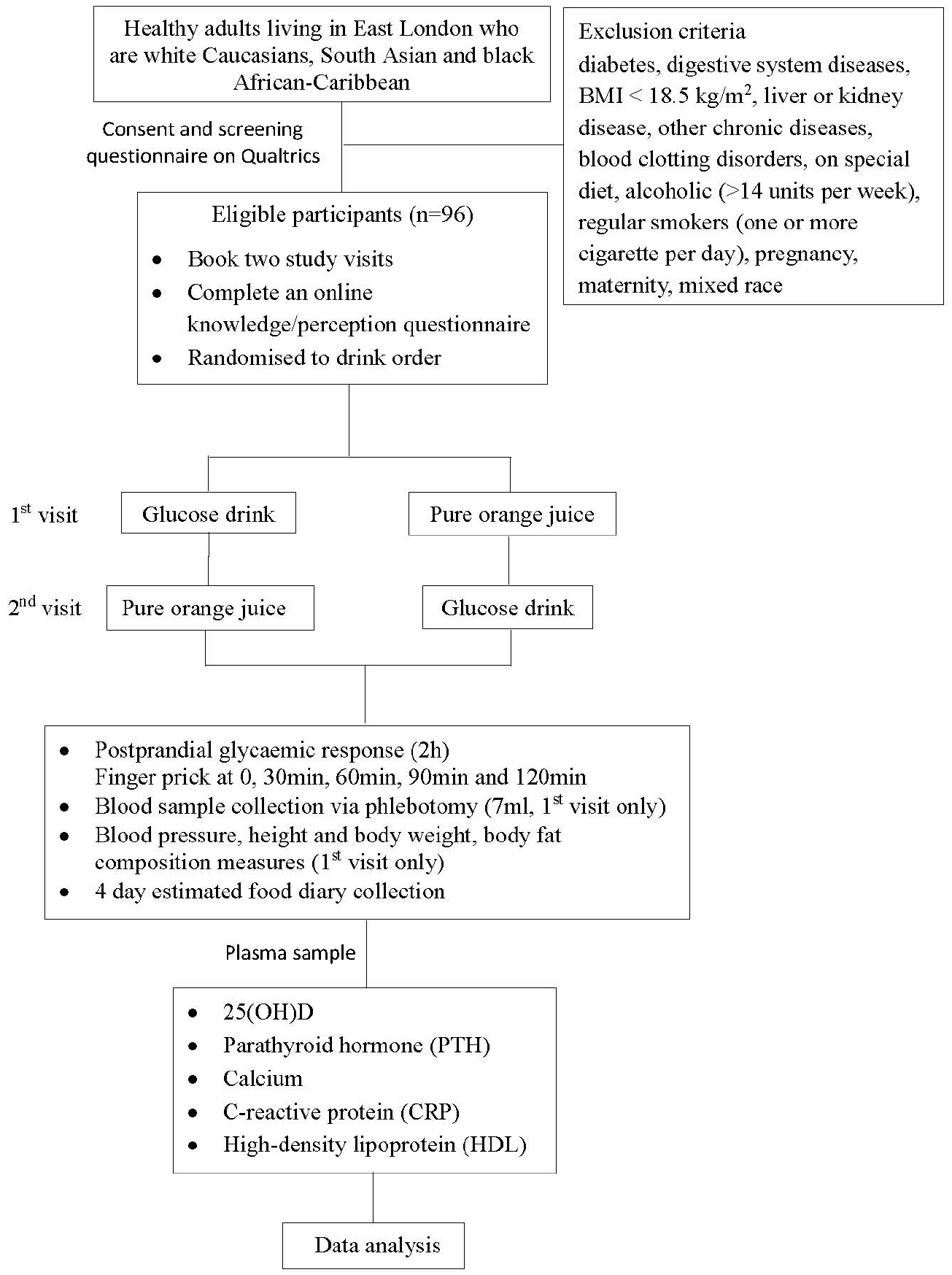
Study flow diagram.

Study status: A) participant recruitment will be completed at the end of December 2024; B) data collection will be completed at the end of December 2024 C) results are expected in January and February. None of these stages have already been completed.

### Participants

The inclusion criteria are as follows: 18–65 y in general good health and living in East London from white, SA or AC origins. The exclusion criteria are as follows: diabetes; digestive system diseases; BMI < 18.5 kg/m2; liver or kidney disease; other chronic diseases; blood clotting disorders; following a special diet; alcohol consumption (>14 units per week); regular smoking (one or more cigarettes per day); pregnancy; maternity; and mixed race. A health and lifestyle questionnaire will be used to screen the eligibility of the participants. Participants will provide informed consent online before being screened for eligibility. Eligible participants will be asked to book their two visits and will receive a shopping voucher worth £20 per visit.

### Recruitment

There are a few methods of participant recruitment. We will recruit staff and students who live in East London with gatekeeper permissions from the Dean of the school. Recruitment adverts will be circulated to staff and students at City, University of London. There are 19975 students, among whom 64% are from the UK, and a large proportion of students are from different London boroughs, including the East London area. Each year, many staff or student projects recruit participants successfully in this way. From local communities in East London with gatekeeper permissions. We will contact local ethnic communities, including the Bangladeshi Community, London Central Mosque, Bangladesh Embassy and Indian & Bangladesh Hindu Community East London, etc. We will also recruit participants via social media, including Meta and Instagram.

### Treatments

The participants will consume a glucose drink (75 g glucose in 300 ml water, 281 kcal) used for the OGTT and pure OJ (Tesco 100% Pure Squeezed Orange Juice Smooth 300 ml containing 129 kcal, 30 g sugar, 0.3 g fibre, 1.8 g protein and 90 mg vitamin C) on separate occasions with at least 48-hour interval and at random order. The two drinks were chosen rather than meals because of fewer facilities needed to cater to participants, less potential food hygiene issues, and being more acceptable to participants from different ethnic backgrounds. The participants fast for 8–12 h. The blood glucose concentration is measured via a HemoCue Glucose 201+ Analyser (Health-care Equipment & Supplies, Surrey UK) at 0, 30, 60, 90 and 120 min before and after drink consumption by finger prick. Two ml of fasting blood will be collected via phlebotomy at the first or second visit only. The plasma is separated by centrifuging the blood sample at 2000 × g for 10 min and stored at -20°C until analysis of some relevant biometabolic parameter measures

During the 2-h study period, the participants are asked to stay sedentary, not eat or drink anything. At the night prior to their study visit, participants are encouraged to follow their normal diet, have good sleep during the night and avoid alcohol and intensive exercise.

### Randomisation

The order of drink consumption is randomised by using the Excel RAND function. In the Excel spreadsheet, a list of participants (from 1-96) is shown in one column. In the next column, we use the RANDBETWEEN function and choose 0 and 1 as the ranges to randomly generate values of 0 or 1. Participants with a value of 0 will consume glucose drink while participants with a value of 1 will consume pure orange juice as their first drink.

### Outcome measures

The primary outcome will be the postprandial glycaemic response (blood glucose concentrations at the above five time points) measured by HemoCue Glucose 201+ Analyser (Health-care Equipment & Supplies, Surrey UK).

Plasma 25(OH)D is the most commonly used indicator of vitD status. PTH and calcium are closely regulated by 25(OH)D levels^(26)^, whereas 25(OH)D is inversely associated with CRP, indicating an anti-inflammatory property of vitD^(27)^. Cholesterol and HLD also have inverse and positive associations, respectively, with vitD status^(28)^. Body mass index (BMI) and fat composition are used as confounding factors of 25(OH)D^(29)^ and PGR in drinks^(30)^. Therefore, the secondary outcomes and measures include plasma 25(OH)D and PTH tested using AIA-900 immunoassay analyser (Tosoh Bioscience, USA); C-reactive protein (CRP), calcium, total cholesterol and high- density lipoprotein (HDL) tested using Horiba Pentra 400 Biochemistry Analyser (Horiba, Japan). In addition, the body weight and fat composition will be measured by TANITA DC-360 P (Tanita, Amsterdam), and body height by stadiometer. Dietary vitD intake will be analysed by a four-day estimated food diary analysed by Nutritics software (Nutritics Ltd., Dublin). Knowledge and perception of vitD will be assessed by a questionnaire collected via Qualtrics survey platform.

The name, email/mobile (for appointment purposes), sex, age, and ethnicity of the participants are also collected.

## Data analysis

The sample size was calculated by G*Power software (version 3.1.9.7; Heinrich-Heine-Universität Düsseldorf, Düsseldorf, Germany). This study aims to achieve a minimum of 25% variability in the postprandial glycaemic response among three ethnic groups, considering that the response is taken from five different time points with 30 min intervals for each person, and to achieve 80% power in the study, we will need 32 people in each group (n=96 in total) at the 5% level of significance. Continuous data are presented as the means ± SDs. Categorical data are presented as percentages. Two-way repeated-measures ANOVA is used to assess time (5 timepoints) effect (within-subject), between-subject effects (ethnicity, sex, normal weight and overweight/obese) and the effect–time interaction. Continuous variables (e.g., 25(OH)D) etc.) are compared between groups via two-way ANOVA. Dietary vitD intake between groups will be analysed by one-way ANOVA (three groups) or an independent t test (two groups) if the data are normally distributed; otherwise, the Mann-Whitney test will be used. Categorical variables, e.g., the percentage of patients with vitD deficiency are compared between groups via Chi-square tests. Data normality will be tested by the Kolmogorov–Smirnov test. The generalized linear mixed models (GLMMs) for longitudinal data will also be used for modelling glycaemic response over time because this model accounts for the correlation between observations within each individual and adjusts for confounding factors (e.g., BMI and age). The statistical significance will be reported with a p value and a 95% confidence interval at the 5% significance level. The statistical software IBM SPSS 29 will be used to analyse the data. The percentage of missing data was low on the basis of our previous experience. Therefore, regression imputation is used to address missing data. Both intention-to-treat analysis and per protocol analysis will be used for the data analysis.

## Ethics and expected outcomes/outputs

The study complies with the Helsinki Declaration II and was approved by the Senate Research Ethics Committee at City, University of London (ETH2223-2000). The findings of the study will be communicated to other researchers, clinical professionals and policymakers primarily through publications in peer-reviewed journals, seminars and conferences. A summary leaflet will be produced in plain English and shared with participants, local communities, GP clinics and hospitals. The leaflet will include actions that can be taken by East London residents in their local food environments. The findings will be also disseminated by university newsletters and social media. This study will increase awareness of the health outcomes of VitD deficiency, particularly in relation to T2DM, and provide rationales to inform education programs and food fortification to combat VitD deficiency in minorities in East London and the wider public. We anticipate that a major outcome of this study will be evidence to inform a randomised controlled trial (RCT) to confirm the causal relationship between vitD status and glycaemic control in ethnic minorities in the UK. Currently, research on vitD supplementation and PGR interventions has produced inconclusive results, and research on minority groups and young adults is needed^(31)^.

## Dissemination plan

- Lay summary reports/leaflets, which will be made available to participants and local communities, and promoted via a social media run by City, University of London (Facebook, Twitter etc.) and University newsletter
- Present the findings in seminars at school, and university level open to professionals or staff and students from other disciplines (e.g., an annual event called Develop@City which focuses on the main themes of Creativity, Wellbeing, Development and Community).
- Professional reports will be made available to professionals at City, University of London, wider partners of Barts Charity, and via LinkedIn
- An abstract of the findings will be submitted to the annual Nutrition Society Conference, or Diabetes UK Professional Conference (organised by Diabetes UK) to share the results with broader nutrition and health care professionals. It is hoped the presentation will generate interest amongst attendees and lead to collaborations in working with the applicant team to prepare an application for a larger project grant.
- A research paper will be produced and submitted to peer review journals that have agreement with City, University of London for publication fee waiver.

## Data Availability

No datasets were generated or analysed during the current study. All relevant data from this study will be made available upon study completion.

## Contributors

HD is the principal investigator of the study and led the design of the study and the preparation of this manuscript and applied for ethics approval. ST contributed to creation of the questionnaire on Qualtrics, and the recruitment advert, CR and SS contributed to the development of the study protocol. SI contributed to the sample size calculation and statistical methods. All authors contributed to the preparation of this manuscript and approved the final manuscript. The funder played no role in the preparation of this manuscript.

## Funding

This work is supported by a Barts Charity Research Seed Grant (Grant Reference Number: G-002602).

There are no declared competing interests.

## Notes

### Competing Interest Statement

The authors have declared no competing interest.

### Clinical Trial

Clinicaltrials.gov ID: NCT06241976

### Funding Statement

Yes

### Author Declarations

This study has been approved by the Senate Ethics Committee at City, University of London (ETH2223-2000).

